# White Matter Alterations in Borderline Personality Disorder

**DOI:** 10.1101/19011676

**Authors:** Isaac Kelleher-Unger, Gabriella Chittano, Zuzanna Tajchman, Iris Vilares

## Abstract

Borderline personality disorder (BorPD) is characterized by instability and impulsivity of mood, relationships and self-image. This disease is an important area of public health policy; compared to other psychiatric disorders, individuals with BorPD experience the most severe functional impairments. Nevertheless, for the patients that do recover, this recovery is stable and only few relapse back to psychopathology. Given its high rate of remission, the rewards of effective treatment options are clear. Identification of underlying anatomical and physiological changes is crucial to refine current treatments and develop new ones. In this perspective, previous magnetic resonance imaging studies have highlighted alterations associated with BorPD phenotype. In particular, diffusion weighted imaging (DWI) has identified many white matter structural alterations in individuals with this diagnosis. Although in its infancy, limiting this line of investigation is a lack of direction at the field level. Hence, the present paper aims to conduct a meta-analysis of DWI findings in individuals with a diagnosis of BorPD, testing the hypothesis that there are specific white matter alterations associated with BorPD. To this end, we performed a meta-analysis of the existing literature of DWI in BorPD representing a total of 123 individuals with BorPD and 117 Controls. Our results indicated that individuals with BorPD show regions of reduced fractional anisotropy in the corpus callosum and fornix. These results survived all jack-knife reshuffles and showed no publication bias. This suggest that alterations in these structures may contribute to psychopathology. Further, the present results lend support to extant psychological and biological models of BorPD.

## Introduction

The Diagnostic and Statistical Manual of Mental Disorders 5 (DSM-5; American Psychological Association) classifies Borderline Personality Disorder (BorPD) as a cluster B Personality disorder. Main characteristics of an individual with BorPD are impulsivity of mood, unstable relationships and distorted self-image.

Mirroring this pattern, the World Health Organisation’s, International Classification of Diseases 10, defines the criteria for Emotionally Unstable Personality Disorder. This disorder is, in concept, like that of BorPD albeit has two sub categories: impulsive and borderline types (ICD-10, World Health Organisation). In common with DSM-5 descriptions, typical behaviours of this disorder include impulsivity, unpredictable mood, and disturbances of self-image – especially in the borderline subtype. Further, and of particular concern, regardless of diagnosing criteria used this disease has an intimate connection with suicidal and self-mutilating practices or actual suicide and self-mutilation (DSM-5, American Psychological Association; ICD-10, World Health Organisation).

Estimates of the global prevalence of BorPD show great variation (Table 1). The biggest studies though estimate a prevalence between 2.7-5.9%, in the general population (Trull et al., 2010; Stinson et al., 2008).

**Table 1.** Reported prevalence of Borderline Personality Disorder amongst different cohorts.

| STUDY | SAMPLE LOCATION | ASSESSMENT | BorPD PREVALANCE (%) |
| --- | --- | --- | --- |
| Torgersen et al. (2001) | Oslo | SIDP-R | 0.7% |
| Samuels et al. (2002) | Baltimore | IPDE | 0.5% |
| Lewin et al. (2005) | NSMHWB | IDPE | 9.8% |
| Coid et al. (2006) | British National Survey | SCID-II | 0.7% |
| Lenzenweger et al. (2007) | NCS-R | IDPE | 1.6% |
| Grant et al. (2008) | NESARC | AUDADIS-IV | 5.9% |
| Trull et al. (2010) | NESARC | AUDADIS-IV | 2.7% |
| Dereboy et al. (2014) | Aydin | DIP-Q | 13.5-15% |

Abbreviations | **NSMHWB**: National Survey of Mental Health and Well-Being, **NCS-R**: National Comorbidity Survey Replication, **NESARC**: National Epidemiological Survey on Conditions Related to Alcohol, **SIDP-R**: Structured Interview for Disorders of Personality-Revised, **IDPE**: International Personality Disorder Examination, **SCID**: Structured Clinical Interview for DSM-IV, **AUDADIS**: The Alcohol Use and Associated Disabilities Interview Schedule, **DIP-Q**: DSM-IV and ICD-10 Personality Questionnaire

The causal factors to developing BorPD are complex (Lieb et al., 2004). It is clear that childhood maltreatment is a risk factor for this psychopathology. In their cohort, Zanarini et al. (2012) report that 62% of the BPD patients disclosed a history of childhood sexual abuse, and, of those, more than 50% of individuals suffered abuse on a weekly basis for at least one year. Though all forms of childhood maltreatment associate with BorPD, childhood emotional abuse confers particular risk for the development of BorPD pathology (Kuo et al., 2015).

Heritability studies suggest a level of genetic influence in the development of BorPD. Estimates from twin studies suggest the mean heritability of BorPD to be 40% (Amad et al., 2014). Supporting this, a large study of 4403 monozygotic twins and 4425 dizygotic twins estimate the heritability of BorPD to be 45%. In particular, GWAS studies report significant differences on chromosome 5 in the SERINC5 gene (Lubke et al. 2013) and, on a gene level analysis, dihydropyrimidine dehydrogenase and plakophilin-4 (Witt et al., 2017). These findings are particularly exceptional given that these are also implicated in schizophrenia, bipolar disorder, and myelination (Krueger et al., 1997; Ripke et al., 2014; Duan et al., 2014). Nevertheless, extrapolation of the results has to be done with caution, since the study by Lubke et al. (2013) comprised of individuals with borderline features assessed by self-report. Nevertheless, these findings suggest a role for environmental and genetic influences in predisposing an individual to BorPD genesis.

Recent meta-analysis of Voxel-Based Morphometry (VBM) studies has highlighted several regions of abnormality in individuals with BorPD (Schulze et al., 2016). Specifically, the right hippocampus, right inferior frontal gyrus and bilateral middle temporal gyri all exhibited grey matter (GM) volume reductions (Schulze et al., 2016). These regions are of particular note as they are robust, surviving all jackknife resamples (Schulze et al., 2016). Individuals with BorPD exhibited increased GM volume in the right supplementary motor area (10/10 jackknife resamples; Schulze et al., 2016). Adding weight to these findings, another VBM meta-analysis performed around the same time, including essentially the same studies (plus an additional one) noted similar findings (Yang et al., 2016). GM volume reductions surviving all 11 jakckknife reshuffles were evident in the left middle temporal gyrus and right inferior frontal gyrus (Yang et al., 2016). Moreover, individuals highlighted increased GM volumed in the right supplementary motor area (Yang et al., 2016). These alterations are further supported by cortical thickness studies. Boen and colleagues et al. (2014) show reduced cortical thickness of the left inferior frontal gyrus, and bilateral paracentral lobule in the individuals with BorPD. Finally, meta-analysis has revealed significantly smaller bilateral amygdala and hippocampi in adults with BorPD (Nunes et al., 2009). Together, these studies suggest that there are structural grey matter differences between BorPD and controls.

Besides grey matter structural differences, functional differences between BorPD and controls have also been analysed. BorPD is often viewed as a disorder of dysregulated emotional responses. In this light, functional magnetic resonance imaging (fMRI) studies, in many cases, utilise behavioural and psychological paradigms, such as sensitivity, emotion regulation and impulsivity paradigms, to probe the neural correlates associated with BorPD (van Zutphen et al., 2015). Indeed, across these three major classes of paradigms, the most robust finding in individuals with BorPD is amygdala hyperactivity in response to emotionally sensitive stimuli (van Zutphen et al., 2015). Schulze et al. (2016)—performing a meta-analysis of neutral vs. negative emotional stimuli—noted hyperactivation in individuals with BorPD in left posterior cingulate gyrus and left middle temporal gyrus. Both of these regions survived all jackknife reshuffles and did not show significant heterogeneity. Conversely, individuals with BorPD exhibit hypoactivation in the left dorso-lateral prefrontal cortex, right dorsal-lateral/medial prefrontal cortex, and left lingual gyrus (Schulze et al., 2016). Again these regions all survive jackknife reshuffles and confirmed no heterogeneity (Schulze et al., 2016). Further, a frequent finding is hypoactivity, in comparison to controls, in the Anterior Cingulate Cortex (ACC) during emotional regulation paradigms whilst impulsivity paradigms yield little direction—most attributed to the deficiency and diversity of studies (van Zutphen et al., 2015). Overall, though, these studies, which the conclusions above are drawn, require an element of vigilance, given that they have low statistical power (van Zutphen et al., 2015).

In resting state functional MRI (rsfMRI), Visintin et al. (2016) provided similar results. Individuals with BorPD displayed resting state hyperactivity in regions of the medial prefrontal cortex (mPFC), ACC and in the precuneus/posterior cingulate cortex (PCC), and exhibited hypoactivation in the right lateral and inferior temporal gyri and bilateral orbitofrontal cortices (Visintin et al., 2016). Having said that, the seven studies included into their meta-analysis were significantly heterogeneous. Other researchers have criticised the paper selection by Visintin et al. (2016). Specifically, Amad and Radua (2017) questioned the inclusion of Soloff et al. (2005) and exclusion of Juengling et al. (2003). The criticisms in large relate to the medication status of the participants in each study, respectively (Amad & Radua, 2017). Therefore, Amad and Radua (2017) replicated the meta-analysis with the altered study list. Agreeing with Visintin et al. (2016), Amad and Radua (2017) noted that individuals with BorPD displayed increased resting state activity in the ACC and in the left inferior and superior frontal gyri compared to controls (Amad & Radua, 2017). However, and contrasting with the results of Visintin et al. (2016), they found that individuals with BorPD showed hypoactivity in the right PCC/precuneus (Amad & Radua, 2017).

From the above, it is clear, even on trend level, that subcortical and frontal regions play a role in BorPD. From this perspective, as noted in fMRI studies, clinical symptoms may arise from functional alterations. Functional alterations and structural alterations are, by nature, linked (Schulze et al., 2016). In addition to functional alternations and grey matter volume differences, impairment of white matter (WM) structures has been demonstrated to affect cognition and behaviour (O’Doherty et al., 2017). Indeed, similar cluster B personality disorder—including: antisocial (Jiang et al., 2017), schizotypal (Yueji et al., 2016) and narcissistic (Nenadic et al., 2015) personality disorders—exhibit WM abnormalities. Given its potential efficaciousness on cognition and behaviour, it is important to analyse if WM abnormalities are also present in the individual with BorPD.

WM structures are studied using diffusion-weighted MRI, also called diffusion-weighted imaging (DWI). DWI gives image contrast based on the differential diffusion of water molecules inside the brain (Huisman, 2010). Diffusion tensor imaging (DTI) decomposes diffusion images into a set of tensors to model the shape of this diffusion (Huisman, 2010), *i.e*. DTI is a particular way of modelling the DWI data (Soares et al., 2013). From this data, one can then calculate the degree of Fractional Anisotropy (FA) of a particular tissue, which gives us information about white matter tracts (Huisman, 2010). The idea behind it is that in white matter tracts the diffusion of water is made primarily over the main axis of the fiber, and much less so in the other directions, i.e. the diffusion is very anisotropic (high FA, appears bright in the image). By contrast, diffusion of water in the cerebrospinal fluid (CSF) is done more homogeneously across all directions, i.e. the diffusion is isotropic (low FA, appears dark in the image) (Huisman, 2010). Based on this, many people have been using DWI/DTI and FA measures to explore the white matter architecture in living humans, including differences in connectivity of the brain areas, white matter fiber orientation and density, as well as myelination (Huisman, 2010; Carrasco et al., 2012; Soares et al., 2013).

Several studies have sought to understand WM abnormalities in BorPD using DWI/DTI (e.g. Grant et al., 2007; Carrasco et al., 2012; Ninomiya et al., 2018). This line of investigation shows promise. Nonetheless, the extant literature has not been discerned as a whole. Indeed, to date (and to our knowledge), no meta-analysis or systematic review of DTI alterations in BorPD exists. In consequence, the principle aim of this study was to scrutinize, at a meta-analysis level, FA changes in individuals with BorPD.

As expressed above, VBM and rs-/fMRI research pointed to the involvement of the subcortical and frontal regions in the psychopathology of BorPD. Accordingly, if any WM alterations exist, they should be in structures connecting these regions. In this view, the principle WM tracts connecting subcortical structures to cortical regions include the superior longitudinal fasciculus, uncinate fasciculus and cingulum bundle (O’Doherty et al., 2017; Schmahmann & Pandya, 2009). Hence, this study aims to test the hypothesis that individuals diagnosed with BorPD will show reduced FA in these structures.

## Methods

The current study was conducted, in large, according to the guidelines set out by Müller et al. (2018). Data sharing is not applicable to this article as no new data were created or analysed in this study.

### Literature Search

Literature search was conducted in April 2018. This search included the PubMed and Web of Science databases with the search terms: (i) borderline personality disorder, (ii) BPD, (iii) diffusion tensor, (iv) diffusion weighted, (v) white matter, and (vi) DTI. Further, combining these keywords produced the string:

> (((borderline personality disorder) OR BPD)) AND ((((diffusion tensor) OR diffusion weighted) OR white matter) OR DTI)

The above search retrieved 181 titles (see Figure 1). At this stage, studies were selected for full text reading if: (i) they were not a duplicate across databases, and (ii) they examined BorPD cohorts. Although a coarse first pass filter, this method proved effective due to the large overlap across databases and the common use of the abbreviation BPD. Indeed, it is worth noting that BPD is common in the extant literature to abbreviate borderline personality disorder, bipolar disorder and bronchopulmonary dysplasia.

**Figure 1.**
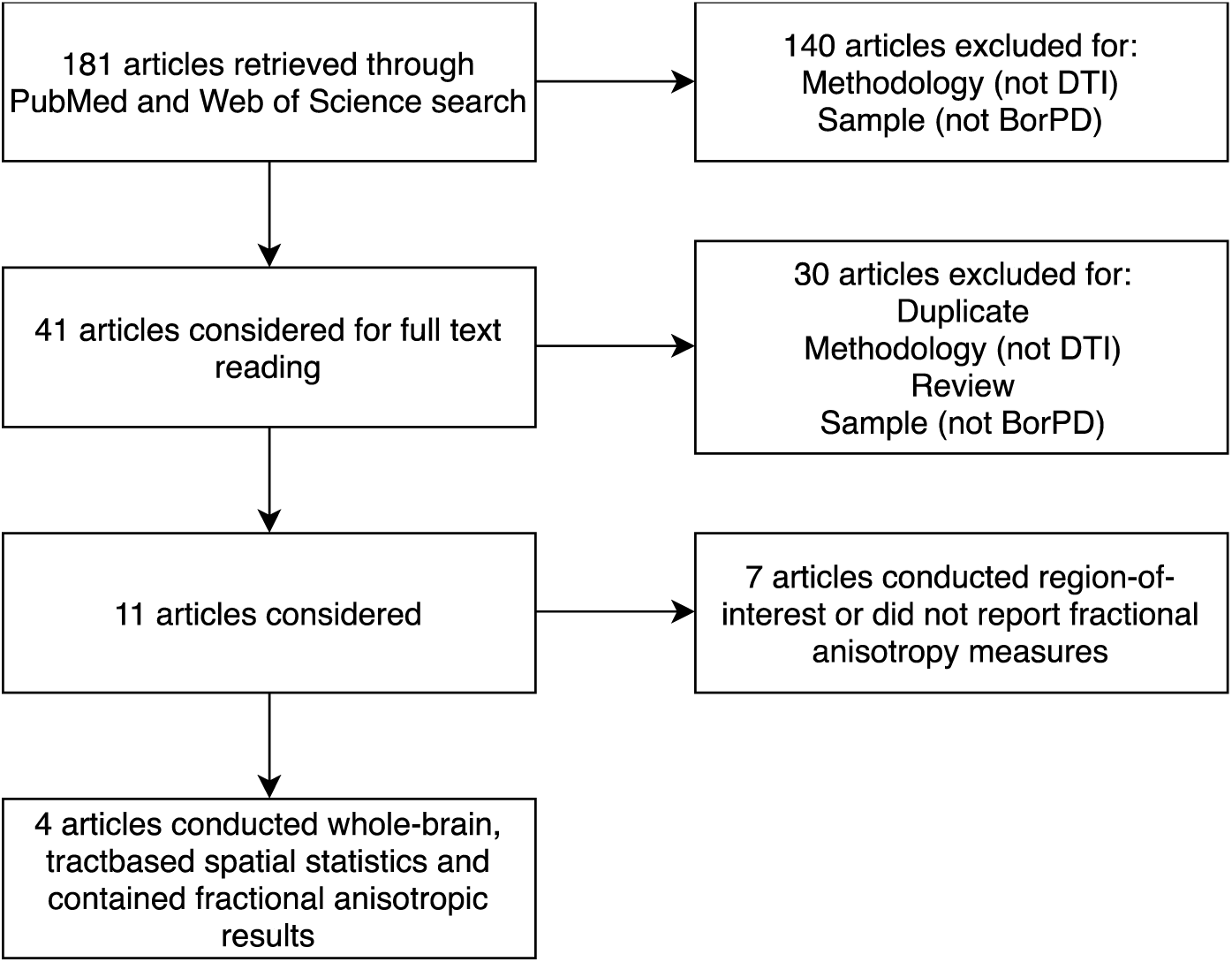
Meta-analysis article selection flowchart.

This first pass identified 41 articles to be considered for full text reading.

Articles read in full were rejected if: (i) the article was not an original article i.e. review, opinion, etc. (ii) the article was a duplicate (iii) the article utilised methodology other than DTI e.g. fMRI (iv) the article studied non-BorPD cohorts. This final filtration left 11 articles for consideration (Carrasco et al., 2012; Gan et al., 2016; Grant et al., 2007; Lischke et al., 2015; Lischke et al., 2017; Maier-Hein et al., 2014; New et al., 2013; Rusch et al., 2007; Salvador et al., 2016; Whalley et al., 2015; Ninomiya et al., 2018).

Of the 11 articles matching the inclusion criteria, 4 of these articles (Carrasco et al., 2012; Gan et al., 2016; Salvador et al., 2016; Whalley et al., 2015) thereafter matched the inclusion criteria for meta-analysis. That is, 4 articles conducted whole brain, Tract-Based Spatial Statistics (TBSS) and contained FA results.

Note: We contacted the authors of the 7 articles that conducted region-of-interest or did not report fractional anisotropy measures in order to obtain the FA values. However, 6 of the authors did not reply (even after multiple attempts), and the one that did reply did not have the data available. Hence, only the 4 articles that contained FA results were included.

### Statistical Tests

We conducted meta-analysis of the whole brain, TBSS, FA alterations between healthy controls (HC) and individuals with BorPD. Meta-analysis was conducted using Seed-Based D Mapping (SDM v. 5.12; Radua & Mataix-Cols, 2012). Hence, where articles reported p or z-values, these were first converted into t-values using an online tool (http://www.sdmproject.com/utilities/?show=Statistics). Subsequently, standardised SDM operations were performed. Pre-processing utilised a TBSS template (included with SDM v. 5.12; Radua et al., 2011; Peters et al., 2012) with a fractional anisotropy correlational template. Moreover, pre-processing used an anisotropy value of 1, Isotropic Full-Width at Half-Maximum FWHM) = 20mm, TBSS mask, and permutation testing with 500 randomisations.

Next, the weighted mean difference of regional TBSS FA results was calculated with the jack-knife reshufflings. Mean differences were then thresholded at a previously optimised level (Muller et al., 2018; Radua et al., 2012) of *p* = 0.005, peak height threshold = 1, and extent threshold = 10. Finally, to examine heterogeneity amongst articles, visual estimates and Ebber tests were performed.

### Power Analysis

Post-hoc power analysis was conducted using the software: G*Power (version 3.1.9.3; http://www.gpower.hhu.de/en.html; Faul et al., 2007; Faul et al., 2009). As this was a post hoc analysis, the one-tailed power was calculated with α=0.005 to match the threshold used in SDM.

## Results

### Study Characteristics

Demographic data are displayed in Table 2; 4 studies matched the inclusion criteria. Together, these studies represent 123 individuals with BorPD and 117 HCs.; within the 123 individuals with BorPD, 91 were female and 32 were male. This is, in comparison to the HC individuals wherein there were 92 female participants and 25 males. Across the 4 studies, the mean age of individuals with BorPD was 28.74 years old and 28.71 years old for the HC group. Overall, studies utilised mixed participants with regards to comorbid psychological diagnoses and medication use. Of particular interest, only one study (Whalley et al., 2015) included participants with comorbid psychological diagnoses.

**Table 2.**
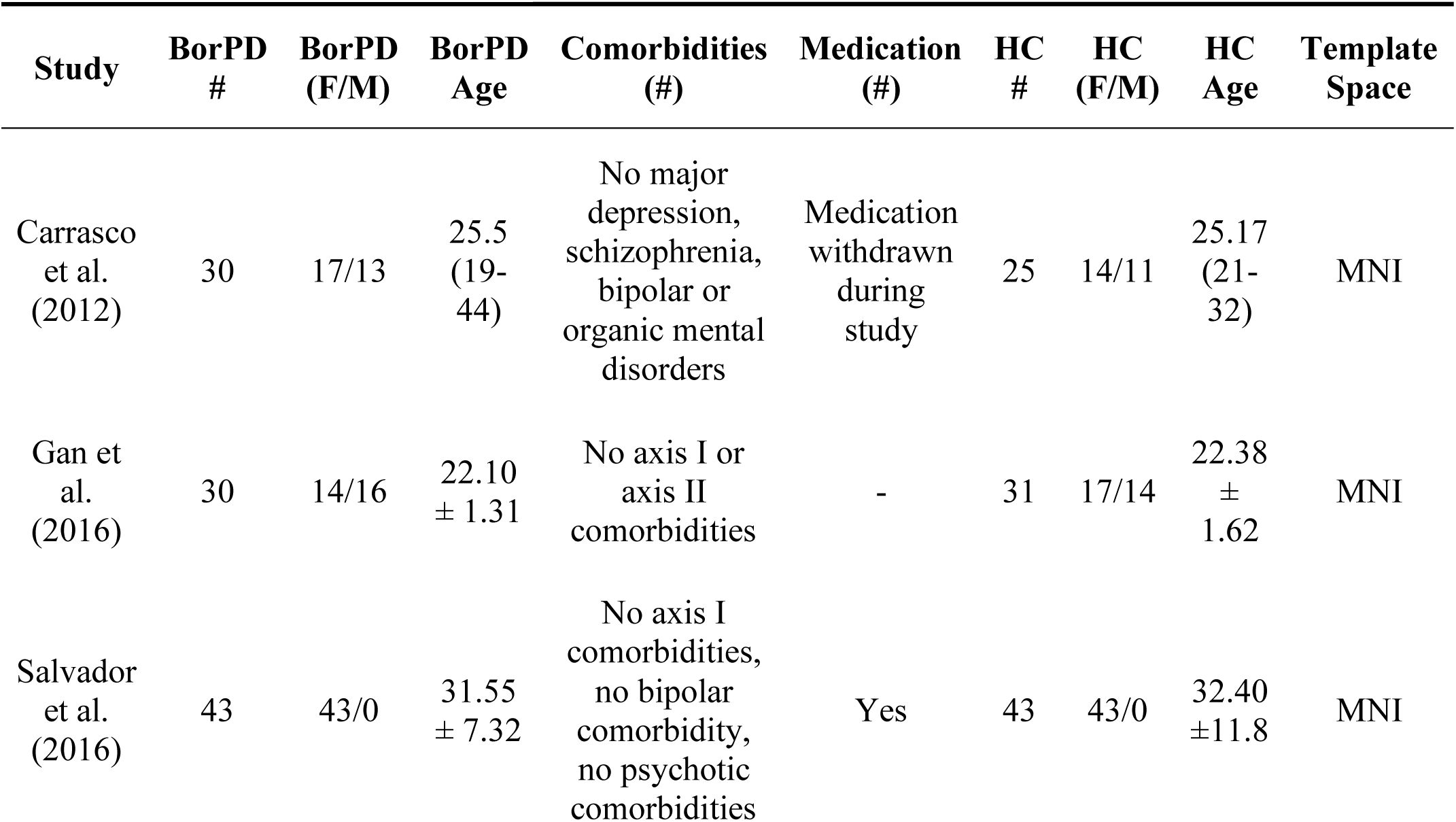
Demographic characteristics of included studies.

### Meta-Analysis

Results of meta-analysis are presented in Figure 2 and Table 3. Results highlighted two clusters of FA reductions. The first foci of reduction, located in the corpus callosum, peaked at MNI coordinates: −16, 22, 22 (Fig. 2, SDM-Z = −1.473, Voxels = 248, p = 0.00007). Further, this result survived all jack-knife reshuffles and showed no publication bias (Fig. 3, bias = −0.07, t = −0.02, p = 0.986). A second peak of reduction was seen in the fornix at MNI coordinates: 0, −6, −16 (Fig. 2, SDM-Z = −1.368, Voxels = 49, p = 0.0001). Similar to the first result, this cluster survived all jack-knife reshuffles and showed no publication bias (Fig. 3, bias = −5.28, t = −1.97, p = 0.188)

**Table 3.**
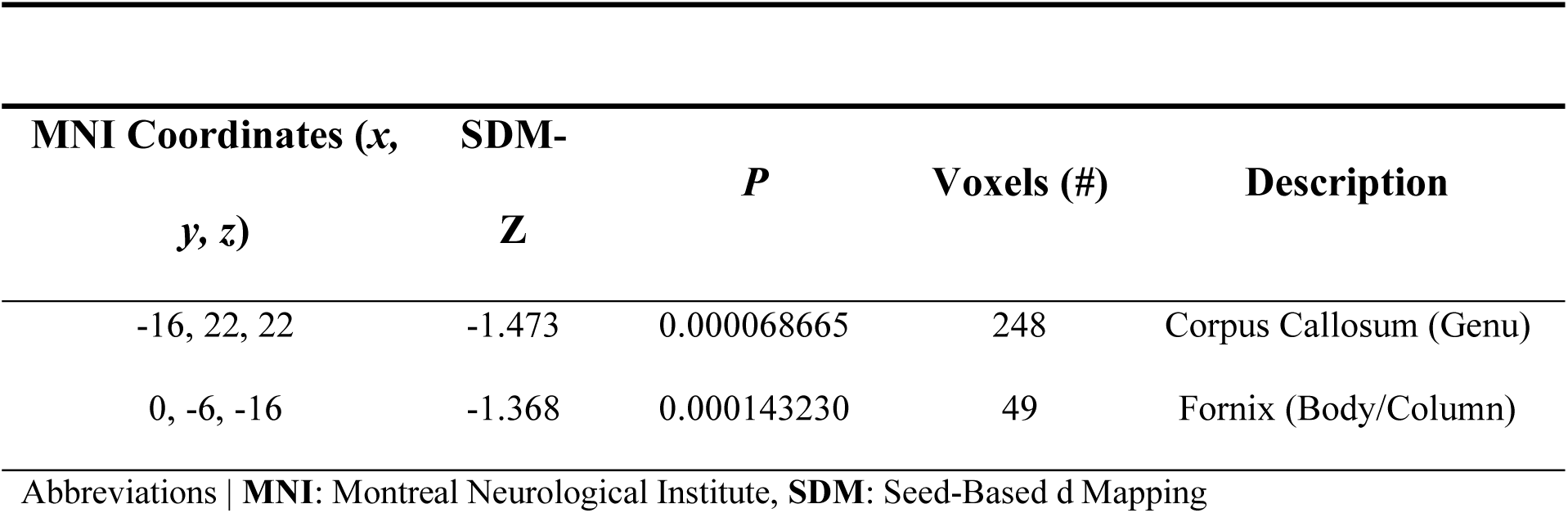
Result of meta-analysis

**Figure 2.**
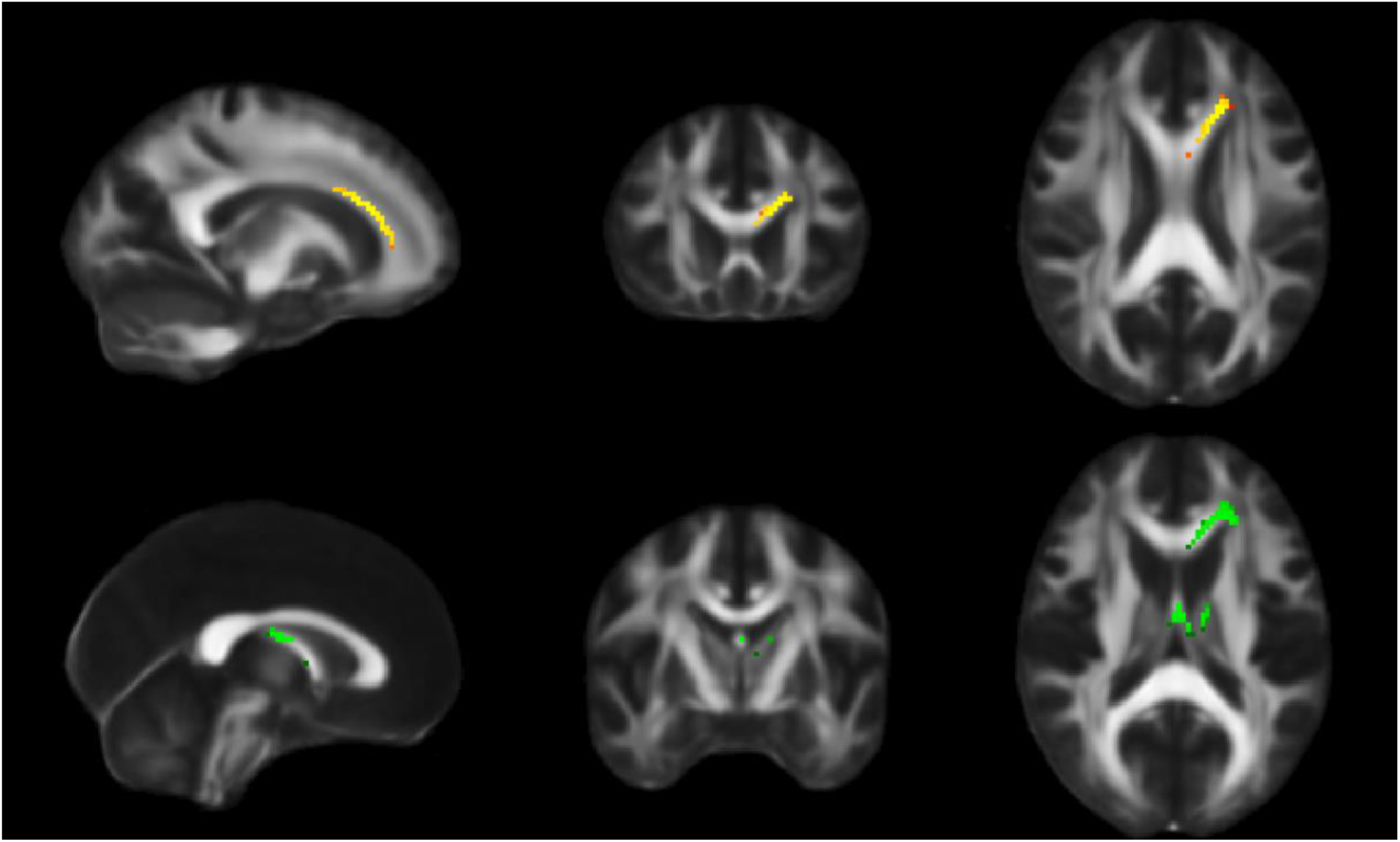
Results of meta-analysis. Meta-analysis was conducted in Seed-based D Mapping using standardised operations. Pre-processing used a tract-based spatial statistics template and a fractional anisotropy correlational template with an anisotropic value of 1 and smoothing kernel of 20mm full width at half maximum. Meta-analysis indicated two foci of significant reductions. (Top) Individuals with borderline personality disorder show reduced fractional anisotropy (SDM-Z = −1.473; p = 0.000068665) in the corpus callosum (MNI: −16, 22, 22; voxels = 248) compared to control. (Bottom) Individuals with borderline personality disorder show reduced fractional anisotropy (SDM-Z = −1.368; p = 0.00143230) in the fornix (MNI: 0, −6, −16; voxels = 49) compared to control.

**Figure 3.**
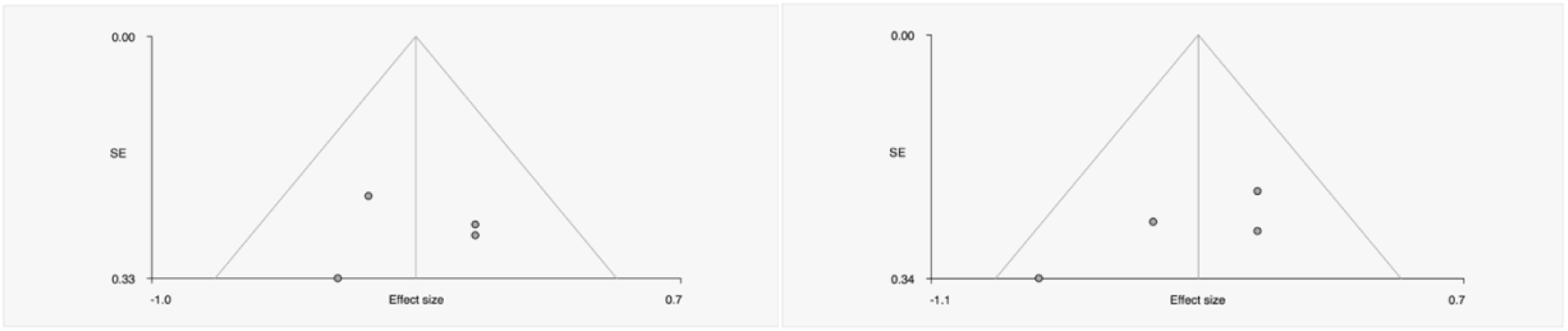
Funnel plots of publication bias. (Left) MNI coordinates: −16, 22, 22 (Right) MNI coordinates: 0, −6, −16. These funnel plot of the 4 studies (Carrasco et al., 2012; Gan et al., 2016; Salvador et al., 2016; Whalley et al., 2015) included in the meta-analysis were produced using the Egger test. The symmetry of the plots suggests an absence of publication bias. This confirmed by the results of the Egger test for both clusters. (Left) bias = −0.07, t = −0.02, p = 0.986 (Right) MNI bias = −5.28, t = −1.97, p = 0.188

### Power Analysis

The results of the post hoc power analysis are presented in Figure 4. The results suggest that with the present sample (n=240), and α=0.005, meta-analysis results were sufficient to detect an effect of d= 0.5 with a power of 0.849.

**Figure 4.**
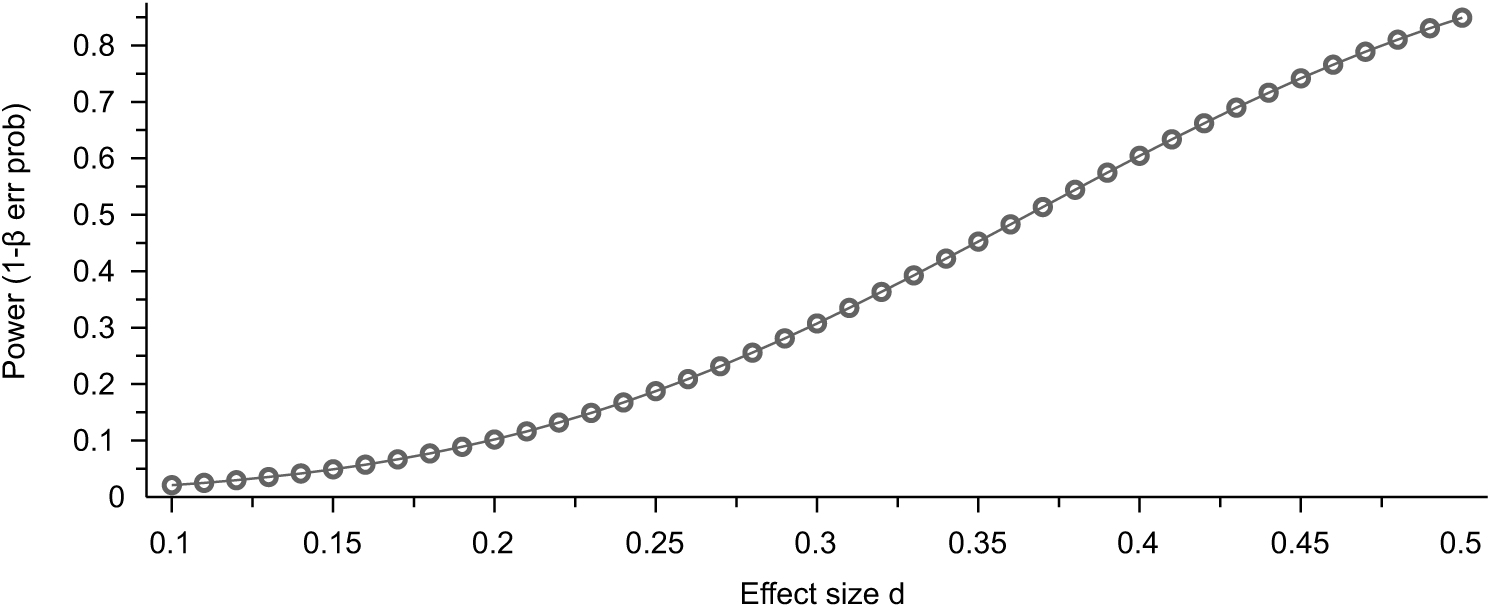
Plot of power vs. effect size. Post hoc power analysis of meta-analysis. Figure highlights the power for various effect sizes for 2-tail, α=0.005, n=240.

**Figure 5.**
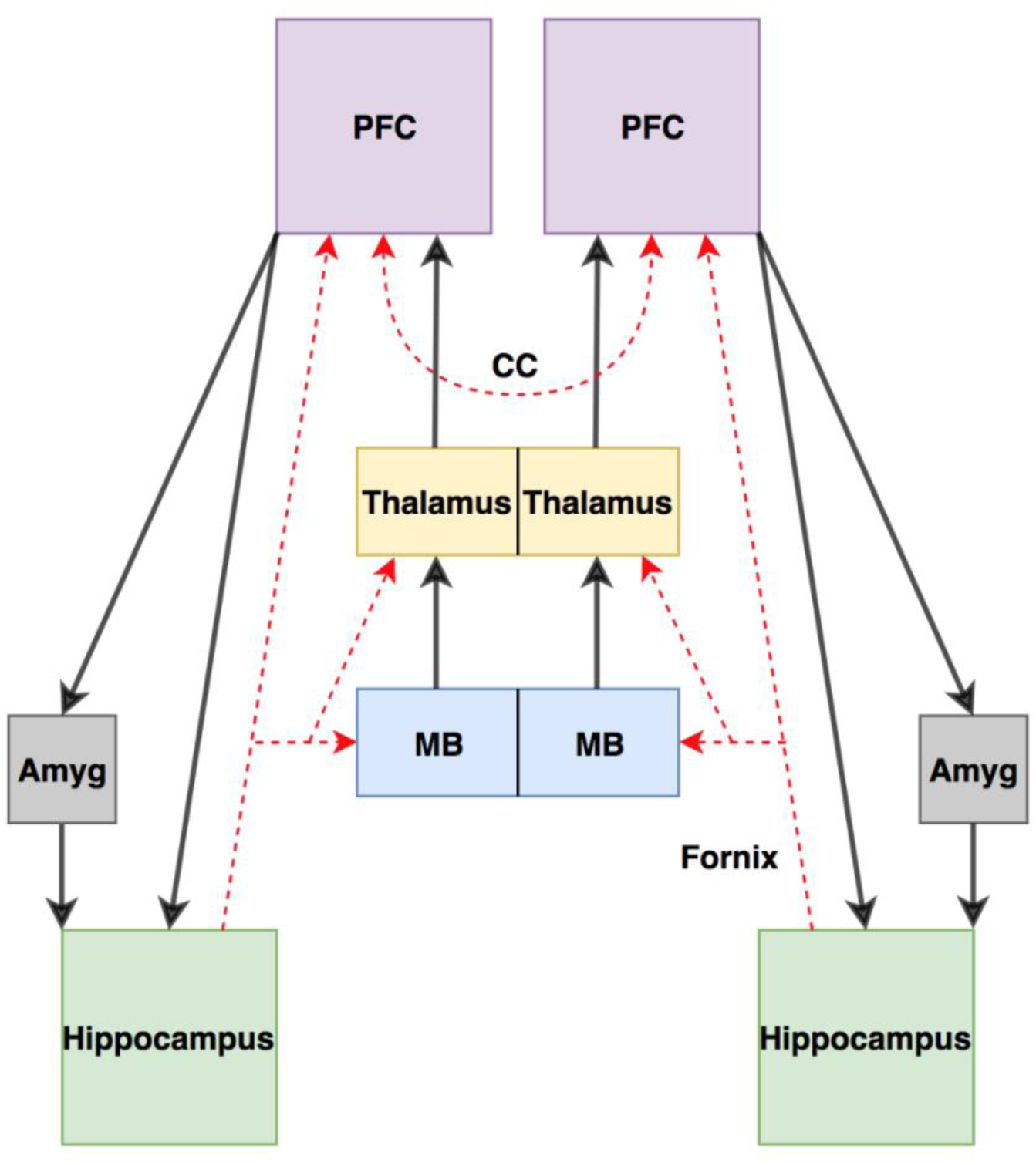
Proposed model of borderline personality disorder. In light of the present results, we propose that white matter alterations in the corpus callosum and fornix may contribute to psychopathology. In particular, abnormal hippocampal/prefrontal connectivity via the fornix may underscore many of the behavioural characteristics of the disorder. Exacerbating this are reductions in corpus callosum integrity. These reductions diminish interhemispheric connectivity and may further contribute to borderline personality disorder behaviours.

## Discussion

The aim of the present study was to conduct a meta-analysis of fractional anisotropic alterations in BorPD. Hence we hypothesised that FA alterations would be noted in the superior longitudinal fasciculus, uncinate fasciculus, and cingulum bundle. Instead, the results of the present study suggest a significant decrease in FA values in the corpus callosum (CC) and fornix. Owing to this result, we reject our original hypothesis.

The CC is the major interhemispheric pathway in the brain (Schmahmann and Pandya, 2009). Main current theories state that its main function is to integrate and facilitate communication between the cerebral hemispheres, although it is not yet clear if this is done through excitatory or inhibitory effects (Toyama et al., 1974; Karayannis et al., 2007; van der Knaap & van der Ham, 2011). However, there is ample evidence that alterations to the CC might be related with a number of psychopathologies. For example, meta-analysis of TBSS results in major depressive disorder and bipolar disorder suggest reduced FA values in the genu of the CC (Chen et al., 2016; Wise et al., 2016). Additionally, alterations to the genu of the CC have been distinguished in personality disorders, namely the cluster A personality disorder: schizotypal personality disorder (Lener et al., 2014); and the cluster C personality disorder: Obsessive-Compulsive disorder (Radua et al., 2014; but see Fan et al., 2016). Further, and more analogous to BorPD, alterations in CC FA values are noted in another cluster B personality disorder: Anti-Social Personality disorder (Sundram et al., 2012; but see Jiang et al., 2017). Altogether, this suggests an association between differences in white matter structure in the CC and different psychopathologies.

Taking into the consideration the available relevant literature, our results argue for a crucial role of the CC in psychopathology. Moreover, we hypothesise that CC alterations, especially in anterior segments of the CC, may show some specificity for personality disorders. It is crucial to emphasize that we do not claim that alterations in anterior segments of the CC represent a biomarker for BorPD. This exaggeration is twofold: first, we advocate the view that personality disorders are complex disorders, potentially with numerous biomarkers. Second, although our result is strong it remains at odds with some previous literature. In fact, a recent study by Niomiya et al. (2018) noted no FA differences in any region of the brain between healthy controls and individuals with BorPD. One factor that could explain this discrepancy is sampled population differences. In the cohort of individuals with BorPD, 68% were male (Niomiya et al., 2018), while in our study 73% of individuals in the BorPD group were female. Furthermore, the average age on our pooled sample was higher (28.7 years old for both BorPD and Controls) compared to their sample (23.3 and 25.8 years old for BorPD and Controls, respectively). Both age and gender can have an effect on the CC thickness (Luders et al., 2010). In addition, the Niomiya and colleagues (2018) study may have been underpowered (only 35 BorPD individuals included, compared to 123 included in our meta-analysis). Further studies should look at the effects of both gender and age on the CC differences (or lack thereof) between BorPD and Controls.

Our meta-analysis did not include region of interest studies. Hence, we turn our attention to these studies. From the outset, it is important to note that, to the best of our knowledge, no study in the context of BorPD has investigated the CC as a whole. However, two studies (Lischke et al., 2017; New et al., 2013) have investigated differences of the CC in BorPD per subregion of the CC. In this context, Lischke and colleagues (2017) report mild, though significant, FA reductions in those with BorPD in the splenium–but not genu or body–of the CC. Further, New et al. (2013) did not observe any FA differences in forceps major or minor between the BorPD and control groups. A critical appraisal of these studies and the current meta-analysis highlights an important point that may underscore the heterogeneity amongst these results. Lischke et al. (2017), New et al. (2013), and the present meta-analysis investigated different elements of BorPD. Contrasting the wide inclusion criteria of the studies included in the meta-analysis, and by extension the meta-analysis itself, Lischke and colleagues (2017) differentiate between those with BorPD without suicidal behaviour and those with BorPD and suicidal behaviour. Moreover, regardless of suicidal behaviour, individuals with BorPD also had to meet DSM-IV criteria for Attention Deficit Hyperactivity Disorder (Lischke et al., 2017). Finally, this study excluded individuals with BorPD who took antipsychotics (Lischke et al., 2017). Overall then, we agree with the authors that this creates a group of individuals with extraordinary homogeneity (Lischke et al., 2017). However, we question the authors using this as a “representative” sample and suggest that, although homogeneous, differences in the present results with those of Lischke et al. (2017) may stem from their overly specific profiling.

Besides the CC, our meta-analysis also found significant FA differences between BorPDs and controls in the fornix (FOR). The FOR is the principle WM bundle that connects the hippocampus to extra-temporal regions, such as the ventral striatum and the prefrontal cortex (PFC) (Christiansen et al., 2016). An exploratory study by Hayes et al. (2015) suggests that DTI reductions in the FOR correlate to the affective elements of anorexia nervosa, including depressive and anxiety symptoms. Building on this idea, our result, is consistent with extant knowledge of classic affective disorders. For example, FOR alterations are a consistent observation in bipolar disorder (Barysheva et al., 2013; Emsell et al., 2013). Forbye, based on lesion observations, some authors have gone so far as to suggest a causal link between FOR alterations and bipolar disease (Rasmussen et al., 2007). Regardless, this conclusion extends from a single case report and, as such, requires circumspection. Finally, in the case of a major depressive disorder, a large-scale study of 150 participants by Hoogenboom and colleagues (2012) supports a role for FOR alterations. From their study, Hoogenboom et al (2012) show reduced FA values in the medial fornix when comparing individuals with non-remitting major depressive disorder to healthy controls. Particularly related to BorPD, adolescents with depression also exhibit the reduced FOR FA compared to healthy controls (Hoogenboom et al., 2012). In all, this may imply an association between FOR alterations and psychopathology.

The antecedent sections have highlighted the function and pathological aspects of the CC and FOR. Now, we will concentrate on the behavioural consequences of CC/FOR pathology. As mentioned above, the FOR is the principle WM tract between the hippocampus and thalamus/PFC (Christiansen et al. (2016). Further, anterior regions of the CC originate/terminate in regions of the PFC (Zarei et al. (2006). The hippocampus is commonly implicated with the encoding of spatial, temporal, object and behavioural memory (Eichenbaum & Cohen, 2014). The PFC is well known for playing a role in emotional regulation; studies have implicated increased PFC activity in attenuating emotional responses (Hariri et al., 2003), voluntary suppression of sadness (Lévesque et al., 2003), and higher order control of emotional responses (Stein et al., 2007). Given that a characteristic element of BorPD is emotional dysregulation, the alterations in PFC function should be evident. Consistently, numerous studies have identified abnormal PFC function during emotional tasks in the ventromedial PFC (Silbersweig et al., 2007), left dorsolateral PFC (Dudas et al., 2017) and bilateral dorsolateral PFC (Ruocco et al., 2013). This might suggest that disrupted connectivity of the CC and FOR may underscore the disrupted functioning of these regions.

In support of this, suggestions for similar schema exist in other disorders with elements of emotional instability and dysregulation. For example, discriminating the Post-Traumatic Stress Disorder (PTSD) is a pattern of hyperarousal (DSM-5, American Psychological Association) and an inability to inhibit fear and emotional responses (O’Doherty et al., 2017). Recent findings revealed FA reductions in the genu of the CC, in individuals with PTSD, that are more severe than in trauma exposed and healthy individuals, respectively (O’Doherty et al., 2017). Moreover, FA reductions in the genu of the CC correlate with grey matter reductions in the rostral anterior cingulate gyrus (O’Doherty et al., 2017). The parallels between BorPD and PTSD extend to resting state functional magnetic resonance imaging findings. Meta-analysis findings in PTSD report hyperactivity in the ventromedial PFC (PTSD<TEC), and hypoactivity in the dorsomedial PFC (PTSD>NTC; Wang et al., 2016). These regions overlap with regions of altered rsfMRI activity in BorPD (Visintin et al., 2016; Amad & Radua, 2017). In the studies of animal models, Bennet and colleagues (2016) show that the core synaptic circuitry implicated in non-human animal models of PTSD involves the amygdala, hippocampus and PFC. In the context of this circuitry (Bennett et al., 2016), our results fit within this model. Considering these together, it can be proposed that the relationship between the hippocampus and PFC may underscore the common affective aspects of these disorders.

Given that literature presented above, especially the phenomenological overlap and consistency of neuroimaging results, we suggest that the neurocircuitry model for PTSD proposed by Rauch and colleagues (2006) can be extrapolated, and combined, with models of BorPD. Overall then, we propose a model of behavioural dysregulation wherein the emotional aspects of BorPD behaviour originate from a breakdown in hippocampus/PFC circuitry, similar to that proposed by Rauch et al (2006) for PTSD. Further, we hypothesise that the breakdown stems from alterations in the CC and FOR as suggested by our results. This model is attractive as it fits the behavioural aspects of BorPD with extant neurocircuitry models and supports the psychological model of BorPD proposed by Linehan et al. (1993) and Fonagy et al. (1996).

Finally, a history of childhood maltreatment is often reported in by individuals with BorPD. As such, we cannot disregard the influence of childhood maltreatment on our results. On this point, there is little doubt about association between experiencing of any form of childhood abuse and brain alterations (Teicher et al., 2016). For example, children exposed to parental verbal abuse—but no other form of abuse—show increased GM density in the primary auditory cortex (Tomoda et al., 2011). In a similar manner, children who witness inter-parental domestic violence show reduced GM volume and thickness in the primary visual cortex (Tomoda et al., 2012). More specific to the current study, childhood maltreatment shows adverse effects on WM integrity, too. It is interesting that WM alterations also seem to follow a modality specific pattern. Children exposed to inter parental domestic violence show reduced FA in the inferior longitudinal fasciculus (Choi et al., 2012), whereas children exposed to parental verbal abuse show FA reductions in the arcuate fasciculus and, most noteworthy in the context of the present results, the fornix (Choi et al., 2009). Further, CC alterations are one of the most prominent and reliable findings in childhood maltreatment (Teicher et al., 2016). Indeed, children exposed to maltreatment appear to have 17% reduction in CC area compared to non-exposed children (Teicher et al., 2004). Further, children exposed to maltreatment show an 11% reduction in the CC area compared to other children with psychiatric disorders but no history of maltreatment (Teicher et al., 2004). This finding is further reinforced by Paul et al. (2008), who note FA alterations in the genu of the CC in adults with history of early life stress.

With these considerations in mind, it is vital to annotate that the model we have proposed for BorPD, shares many commonalities with the childhood maltreatment model suggested by Teicher and colleagues (2016). Both of these models highlight the importance of the amygdala, hippocampus, prefrontal cortex and fornix. Further, CC alterations are a prime candidate to unveil behaviours seen in BorPD. Teicher and colleagues (2016) suggest that a breakdown of interhemispheric connectivity, resultant from reduced CC integrity, may underscore splitting behaviours in individuals with BorPD.

### Limitations & Conclusion

The present analysis included 4 articles representing 123 individuals with BorPD and 117 control individuals. This is a very small sample size and questions the necessity of a meta-analysis. However we propose that, although a small sample size for a meta-analysis, the presented meta-analysis has much greater statistical power than any of the included studies. Furthermore, the Egger tests of heterogeneity were not significant. i.e. no evidence of publication bias, and the results of the meta-analysis remained significant following jacknife shuffles. This suggest that, whilst the sample size is ungenerous, the results extending from this study are robust and acceptable.

Furthermore, the heterogeneity of the participants within each study deserves mentioning. In a statistical sense, there was no publication bias or heterogeneity amongst the studies included (Fig. 3). Yet, in a demographic sense, Table 2 highlights that there is an element of heterogeneity between individual cohorts. From this perspective, some elements merit particular deliberation: first is gender. In the meta-analysis, of the 123 individuals represented in the BorPD group, 91 (73.9%) were female. There is a long-held view that the ratio of female-to-male BorPD diagnoses is 3:1 (DSM-5, American Psychological Association). The DSM-5 suggests that 75% of individuals with BorPD are female (DSM-5, American Psychological Association). Thus, while gender could be a potential confound to the results, this is debateable. Females have greater odds of a BorPD diagnosis than males (Trull et al., 2010). Further, more females are more keen to seek help with mental health issues than males (Tomko et al., 2014). Hence, it can be argued that this skew is representative of the existing BorPD population.

Finally, an element of this study that limits its impact is that the protocol was not pre-registered. This limitation arise from the recommendations outlined by Müller and colleagues (2018). While we made every effort to conduct a transparent analysis, the study protocol was not previously registered. In the future, under different circumstances, any intended analysis should be pre-planned and registered, in line with gold standard recommendations.

The future direction for this work may take many avenues. Importantly, we must reiterate, this work should not be viewed in the context of biomarkers. Rather, the present results should serve as the basis for future primary investigations. Predominantly though, the results of this study require retesting in a primary investigation. Moreover, this line of exploration will benefit from a large-scale cohort and, in the best case, individuals spanning different levels of symptoms to establish a dose-response relationship. From here, subsequent research can then aim to look for a causal factor underlying this pathology. At this stage, investigators may need to pursue genomic or proteomics. The ultimate objective of this approach is to develop psychological or pharmacological treatments that can restore the observable deficits.

## Conclusion

The results of this meta-analysis, representing 240 individuals, support the conclusion that fractional anisotropic changes in the corpus callosum and fornix may contribute to the psychopathology of BorPD. This is a robust result that integrates with extant psychological and neurobiological literature. These results will now require primary investigation, in which what is presented in the paper can serve as the basis for future hypotheses.

## Data Availability

This is a meta-analysis of existing papers in the literature and all the data used came from that publicly-available data. The papers from which data was extracted from are clearly indicated in the manuscript.

## References

Amad, A., & Radua, J. (2017). Resting-state meta-analysis in Borderline Personality Disorder: Is the fronto-limbic hypothesis still valid?. Journal of affective disorders, 212, 7.

Amad, A., Ramoz, N., Thomas, P., & Gorwood, P. (2016). The age-dependent plasticity highlights the conceptual interface between borderline personality disorder and PTSD. European archives of psychiatry and clinical neuroscience, 266(4), 373–375.

Amad, A., Ramoz, N., Thomas, P., Jardri, R., & Gorwood, P. (2014). Genetics of borderline personality disorder: systematic review and proposal of an integrative model. Neuroscience & Biobehavioral Reviews, 40, 6–19.

American Psychiatric Association. (2013). Diagnostic and statistical manual of mental disorders (DSM-5®). American0020Psychiatric Pub.

Barysheva, M., Jahanshad, N., Foland-Ross, L., Altshuler, L. L., & Thompson, P. M. (2013). White matter microstructural abnormalities in bipolar disorder: a whole brain diffusion tensor imaging study. NeuroImage: clinical, 2, 558–568.

Bennett, M. R., Hatton, S. N., & Lagopoulos, J. (2016). Stress, trauma and PTSD: translational insights into the core synaptic circuitry and its modulation. Brain structure and function, 221(5), 2401–2426.

Bøen, E., Westlye, L. T., Elvsåshagen, T., Hummelen, B., Hol, P. K., Boye, B., … & Malt, U. F. (2014). Regional cortical thinning may be a biological marker for borderline personality disorder. Acta psychiatrica scandinavica, 130(3), 193–204.

Carrasco, J. L., Tajima-Pozo, K., Díaz-Marsá, M., Casado, A., López-Ibor, J. J., Arrazola, J., & Yus, M. (2012). Microstructural white matter damage at orbitofrontal areas in borderline personality disorder. Journal of Affective Disorders, 139(2), 149–153.

Choi, J., Jeong, B., Polcari, A., Rohan, M. L., & Teicher, M. H. (2012). Reduced fractional anisotropy in the visual limbic pathway of young adults witnessing domestic violence in childhood. Neuroimage, 59(2), 1071–1079.

Choi, J., Jeong, B., Rohan, M. L., Polcari, A. M., & Teicher, M. H. (2009). Preliminary evidence for white matter tract abnormalities in young adults exposed to parental verbal abuse. Biological psychiatry, 65(3), 227–234.

Christiansen, K., Aggleton, J. P., Parker, G. D., O’Sullivan, M. J., Vann, S. D., & Metzler-Baddeley, C. (2016). The status of the precommissural and postcommissural fornix in normal ageing and mild cognitive impairment: An MRI tractography study. NeuroImage, 130, 35–47.

Christiansen, K., Dillingham, C. M., Wright, N. F., Saunders, R. C., Vann, S. D., & Aggleton, J. P. (2016). Complementary subicular pathways to the anterior thalamic nuclei and mammillary bodies in the rat and macaque monkey brain. European Journal of Neuroscience, 43(8), 1044–1061.

Ripke, S., Neale, B. M., Corvin, A., Walters, J. T., Farh, K. H., Holmans, P. A., … & Pers, T. H. (2014). Biological insights from 108 schizophrenia-associated genetic loci. Nature, 511(7510), 421.

Dereboy, C., Güzel, H. S., Dereboy, F., Okyay, P., & Eskin, M. (2014). Personality disorders in a community sample in Turkey: Prevalence, associated risk factors, temperament and character dimensions. International Journal of Social Psychiatry, 60(2), 139–147.

Distel, M. A., Middeldorp, C. M., Trull, T. J., Derom, C. A., Willemsen, G., & Boomsma, D. I. (2011). Life events and borderline personality features: the influence of gene–environment interaction and gene–environment correlation. Psychological medicine, 41(4), 849–860.

Dudas, R. B., Mole, T. B., Morris, L. S., Denman, C., Hill, E., Szalma, B., … & Voon, V. (2017). Amygdala and dlPFC abnormalities, with aberrant connectivity and habituation in response to emotional stimuli in females with BPD. Journal of affective disorders, 208, 460–466.

Eichenbaum, H., & Cohen, N. J. (2014). Can we reconcile the declarative memory and spatial navigation views on hippocampal function?. Neuron, 83(4), 764–770.

Fan, S., van den Heuvel, O. A., Cath, D. C., van der Werf, Y. D., de Wit, S. J., de Vries, F. E., … & Pouwels, P. J. (2016). Mild white matter changes in un-medicated obsessive-compulsive disorder patients and their unaffected siblings. Frontiers in neuroscience, 9, 495.

Fonagy, P., & Luyten, P. (2009). A developmental, mentalization-based approach to the understanding and treatment of borderline personality disorder. Development and psychopathology, 21(4), 1355–1381.

Fonagy, P., Luyten, P., Allison, E., & Campbell, C. (2017). What we have changed our minds about: Part 1. Borderline personality disorder as a limitation of resilience. Borderline Personality Disorder and Emotion Dysregulation, 4(1), 11.

Gan, J., Yi, J., Zhong, M., Cao, X., Jin, X., Liu, W., & Zhu, X. (2016). Abnormal white matter structural connectivity in treatment-naïve young adults with borderline personality disorder. Acta Psychiatrica Scandinavica, 134(6), 494–503.

Grant, J. E., Correia, S., Brennan-Krohn, T., Malloy, P. F., Laidlaw, D. H., & Schulz, S. C. (2007). Frontal white matter integrity in borderline personality disorder with self-injurious behavior. The Journal of neuropsychiatry and clinical neurosciences, 19(4), 383–390.

Gross, R., Olfson, M., Gameroff, M., Shea, S., Feder, A., Fuentes, M., … & Weissman, M. M. (2002). Borderline personality disorder in primary care. Archives of Internal Medicine, 162(1), 53–60.

Hariri, A. R., Mattay, V. S., Tessitore, A., Fera, F., & Weinberger, D. R. (2003). Neocortical modulation of the amygdala response to fearful stimuli. Biological psychiatry, 53(6), 494–501.

Hayes, D. J., Lipsman, N., Chen, D. Q., Woodside, D. B., Davis, K. D., Lozano, A. M., & Hodaie, M. (2015). Subcallosal cingulate connectivity in anorexia nervosa patients differs from healthy controls: a multi-tensor tractography study. Brain stimulation, 8(4), 758–768.

Hoogenboom, W. S., Perlis, R. H., Smoller, J. W., Zeng-Treitler, Q., Gainer, V. S., Murphy, S. N., … & Iosifescu, D. V. (2014). Limbic system white matter microstructure and long-term treatment outcome in major depressive disorder: a diffusion tensor imaging study using legacy data. The World Journal of Biological Psychiatry, 15(2), 122–134.

Juengling, F. D., Schmahl, C., Hesslinger, B., Ebert, D., Bremner, J. D., Gostomzyk, J., … & Lieb, K. (2003). Positron emission tomography in female patients with borderline personality disorder. Journal of Psychiatric Research, 37(2), 109–115.

Karayannis, T., Huerta-Ocampo, I., & Capogna, M. (2006). GABAergic and pyramidal neurons of deep cortical layers directly receive and differently integrate callosal input. Cerebral Cortex, 17(5), 1213–1226.

Kuo, J. R., Khoury, J. E., Metcalfe, R., Fitzpatrick, S., & Goodwill, A. (2015). An examination of the relationship between childhood emotional abuse and borderline personality disorder features: The role of difficulties with emotion regulation. Child abuse & neglect, 39, 147–155.

Lener, M. S., Wong, E., Tang, C. Y., Byne, W., Goldstein, K. E., Blair, N. J., … & Rimsky, L. S. (2014). White matter abnormalities in schizophrenia and schizotypal personality disorder. Schizophrenia bulletin, 41(1), 300–310.

Lévesque, J., Eugene, F., Joanette, Y., Paquette, V., Mensour, B., Beaudoin, G., … & Beauregard, M. (2003). Neural circuitry underlying voluntary suppression of sadness. Biological psychiatry, 53(6), 502–510.

Lieb, K., Zanarini, M. C., Schmahl, C., Linehan, M. M., & Bohus, M. (2004). Borderline personality disorder. The Lancet, 364(9432), 453–461.

Linehan, M. M. (1993). Cognitive-behavioral treatment of borderline personality disorder. Guilford Publications.

Lischke, A., Domin, M., Freyberger, H. J., Grabe, H. J., Mentel, R., Bernheim, D., & Lotze, M. (2015). Structural alterations in white-matter tracts connecting (para-) limbic and prefrontal brain regions in borderline personality disorder. Psychological medicine, 45(15), 3171–3180.

Lischke, A., Domin, M., Freyberger, H. J., Grabe, H. J., Mentel, R., Bernheim, D., & Lotze, M. (2017). Structural alterations in the Corpus callosum are associated with suicidal behavior in women with borderline personality disorder. Frontiers in human neuroscience, 11, 196.

Lubke, G. H., Laurin, C., Amin, N., Hottenga, J. J., Willemsen, G. A. H. M., Van Grootheest, G., … & Penninx, B. W. J. H. (2014). Genome-wide analyses of borderline personality features. Molecular psychiatry, 19(8), 923.

Luders, E., Thompson, P. M., & Toga, A. W. (2010). The development of the corpus callosum in the healthy human brain. Journal of Neuroscience, 30(33), 10985–10990.

Maier-Hein, K. H., Brunner, R., Lutz, K., Henze, R., Parzer, P., Feigl, N., … & Stieltjes, B. (2014). Disorder-specific white matter alterations in adolescent borderline personality disorder. Biological psychiatry, 75(1), 81–88.

Müller, V. I., Cieslik, E. C., Laird, A. R., Fox, P. T., Radua, J., Mataix-Cols, D., … & Wager, T. D. (2018). Ten simple rules for neuroimaging meta-analysis. Neuroscience & Biobehavioral Reviews, 84, 151–161.

Nemoda, Z., Lyons-Ruth, K., Szekely, A., Bertha, E., Faludi, G., & Sasvari-Szekely, M. (2010). Association between dopaminergic polymorphisms and borderline personality traits among at-risk young adults and psychiatric inpatients. Behavioral and Brain Functions, 6(1), 4.

Nenadic, I., Güllmar, D., Dietzek, M., Langbein, K., Steinke, J., & Gaser, C. (2015). Brain structure in narcissistic personality disorder: a VBM and DTI pilot study. Psychiatry Research: Neuroimaging, 231(2), 184–186.

New, A. S., Carpenter, D. M., Perez-Rodriguez, M. M., Ripoll, L. H., Avedon, J., Patil, U., … & Goodman, M. (2013). Developmental differences in diffusion tensor imaging parameters in borderline personality disorder. Journal of psychiatric research, 47(8), 1101–1109.

Ninomiya, T., Oshita, H., Kawano, Y., Goto, C., Matsuhashi, M., Masuda, K., … & Kanehisa, M. (2018). Reduced white matter integrity in borderline personality disorder: a diffusion tensor imaging study. Journal of affective disorders, 225, 723–732.

Nunes, P. M., Wenzel, A., Borges, K. T., Porto, C. R., Caminha, R. M., & de Oliveira, I. R. (2009). Volumes of the hippocampus and amygdala in patients with borderline personality disorder: a meta-analysis. Journal of personality disorders, 23(4), 333–345.

Paul, R., Henry, L., Grieve, S. M., Guilmette, T. J., Niaura, R., Bryant, R., … & Gordon, E. (2008). The relationship between early life stress and microstructural integrity of the corpus callosum in a non-clinical population. Neuropsychiatric disease and treatment, 4(1), 193.

Rauch, S. L., Shin, L. M., & Phelps, E. A. (2006). Neurocircuitry models of posttraumatic stress disorder and extinction: human neuroimaging research—past, present, and future. Biological psychiatry, 60(4), 376–382.

Ruocco, A. C., Amirthavasagam, S., Choi-Kain, L. W., & McMain, S. F. (2013). Neural correlates of negative emotionality in borderline personality disorder: an activation-likelihood-estimation meta-analysis. Biological Psychiatry, 73(2), 153–160.

Rüsch, N., Weber, M., Il’yasov, K. A., Lieb, K., Ebert, D., Hennig, J., & van Elst, L. T. (2007). Inferior frontal white matter microstructure and patterns of psychopathology in women with borderline personality disorder and comorbid attention-deficit hyperactivity disorder. Neuroimage, 35(2), 738–747.

Salvador, R., Vega, D., Pascual, J. C., Marco, J., Canales-Rodríguez, E. J., Aguilar, S., … & Maristany, T. (2016). Converging medial frontal resting state and diffusion-based abnormalities in borderline personality disorder. Biological psychiatry, 79(2), 107–116.

Schmahmann, J., & Pandya, D. (2009). Fiber pathways of the brain. OUP USA.

Schulze, L., Schmahl, C., & Niedtfeld, I. (2016). Neural correlates of disturbed emotion processing in borderline personality disorder: a multimodal meta-analysis. Biological psychiatry, 79(2), 97–106.

Silbersweig, D., Clarkin, J. F., Goldstein, M., Kernberg, O. F., Tuescher, O., Levy, K. N., … & Epstein, J. (2007). Failure of frontolimbic inhibitory function in the context of negative emotion in borderline personality disorder. American Journal of Psychiatry, 164(12), 1832–1841.

Soloff, P. H., Price, J. C., Meltzer, C. C., Fabio, A., Frank, G. K., & Kaye, W. H. (2007). 5HT2A receptor binding is increased in borderline personality disorder. Biological psychiatry, 62(6), 580–587.

Soloff, P. H., Abraham, K., Burgess, A., Ramaseshan, K., Chowdury, A., & Diwadkar, V. A. (2017). Impulsivity and aggression mediate regional brain responses in Borderline Personality Disorder: An fMRI study. Psychiatry Research: Neuroimaging, 260, 76–85.

Stein, J. L., Wiedholz, L. M., Bassett, D. S., Weinberger, D. R., Zink, C. F., Mattay, V. S., & Meyer-Lindenberg, A. (2007). A validated network of effective amygdala connectivity. Neuroimage, 36(3), 736–745.

Stinson, F. S., Dawson, D. A., Goldstein, R. B., Chou, S. P., Huang, B., Smith, S. M., … & Grant, B. F. (2008). Prevalence, correlates, disability, and comorbidity of DSM-IV narcissistic personality disorder: results from the wave 2 national epidemiologic survey on alcohol and related conditions. The Journal of clinical psychiatry, 69(7), 1033.

Teicher, M. H., Dumont, N. L., Ito, Y., Vaituzis, C., Giedd, J. N., & Andersen, S. L. (2004). Childhood neglect is associated with reduced corpus callosum area. Biological psychiatry, 56(2), 80–85.

Teicher, M. H., Samson, J. A., Anderson, C. M., & Ohashi, K. (2016). The effects of childhood maltreatment on brain structure, function and connectivity. Nature Reviews Neuroscience, 17(10), 652.

Tomko, R. L., Trull, T. J., Wood, P. K., & Sher, K. J. (2014). Characteristics of borderline personality disorder in a community sample: comorbidity, treatment utilization, and general functioning. Journal of personality disorders, 28(5), 734–750.

Tomoda, A., Polcari, A., Anderson, C. M., & Teicher, M. H. (2012). Reduced visual cortex gray matter volume and thickness in young adults who witnessed domestic violence during childhood. PloS one, 7(12), e52528.

Tomoda, A., Sheu, Y. S., Rabi, K., Suzuki, H., Navalta, C. P., Polcari, A., & Teicher, M. H. (2011). Exposure to parental verbal abuse is associated with increased gray matter volume in superior temporal gyrus. Neuroimage, 54, S280–S286.

Toyama, K., Matsunami, K., Ohno, T., & Tokashiki, S. (1974). An intracellular study of neuronal organization in the visual cortex. Experimental brain research, 21(1), 45–66.

Trull, T. J., Jahng, S., Tomko, R. L., Wood, P. K., & Sher, K. J. (2010). Revised NESARC personality disorder diagnoses: gender, prevalence, and comorbidity with substance dependence disorders. Journal of personality disorders, 24(4), 412–426.

van der Knaap, L. J., & van der Ham, I. J. (2011). How does the corpus callosum mediate interhemispheric transfer? A review. Behavioural brain research, 223(1), 211–221.

van Zutphen, L., Siep, N., Jacob, G. A., Goebel, R., & Arntz, A. (2015). Emotional sensitivity, emotion regulation and impulsivity in borderline personality disorder: a critical review of fMRI studies. Neuroscience & Biobehavioral Reviews, 51, 64–76.

Visintin, E., De Panfilis, C., Amore, M., Balestrieri, M., Wolf, R. C., & Sambataro, F. (2016). Mapping the brain correlates of borderline personality disorder: a functional neuroimaging meta-analysis of resting state studies. Journal of affective disorders, 204, 262–269.

Wang, T., Liu, J., Zhang, J., Zhan, W., Li, L., Wu, M., … & Gong, Q. (2016). Altered resting-state functional activity in posttraumatic stress disorder: A quantitative meta-analysis. Scientific reports, 6, 27131.

Welcome, S. E., & Chiarello, C. (2008). How dynamic is interhemispheric interaction? Effects of task switching on the across-hemisphere advantage. Brain and cognition, 67(1), 69–75.

Whalley, H. C., Nickson, T., Pope, M., Nicol, K., Romaniuk, L., Bastin, M. E., … & Hall, J. (2015). White matter integrity and its association with affective and interpersonal symptoms in borderline personality disorder. NeuroImage: Clinical, 7, 476–481.

Wise, T., Radua, J., Nortje, G., Cleare, A. J., Young, A. H., & Arnone, D. (2016). Voxel-based meta-analytical evidence of structural disconnectivity in major depression and bipolar disorder. Biological psychiatry, 79(4), 293–302.

Witt, S. H., Streit, F., Jungkunz, M., Frank, J., Awasthi, S., Reinbold, C. S., … & Dietl, L. (2017). Genome-wide association study of borderline personality disorder reveals genetic overlap with bipolar disorder, major depression and schizophrenia. Translational psychiatry, 7(6), e1155.

World Health Organization. (1990). International Classification of Diseases (ICD-10).[Internet. Accessed December 17, 2013.]. World Health Organization.

Yang, X., Hu, L., Zeng, J., Tan, Y., & Cheng, B. (2016). Default mode network and frontolimbic gray matter abnormalities in patients with borderline personality disorder: A voxel-based meta-analysis. Scientific reports, 6, 34247.

Zanarini, M. C., Frankenburg, F. R., Reich, D. B., & Fitzmaurice, G. (2012). Attainment and stability of sustained symptomatic remission and recovery among patients with borderline personality disorder and axis II comparison subjects: a 16-year prospective follow-up study. American Journal of Psychiatry, 169(5), 476–483.

Zarei, M., Johansen-Berg, H., Smith, S., Ciccarelli, O., Thompson, A. J., & Matthews, P. M. (2006). Functional anatomy of interhemispheric cortical connections in the human brain. Journal of anatomy, 209(3), 311–320.

Zhang, J., Kendrick, K. M., Lu, G., & Feng, J. (2014). The fault lies on the other side: Altered brain functional connectivity in psychiatric disorders is mainly caused by counterpart regions in the opposite hemisphere. Cerebral Cortex, 25(10), 3475–3486.

